# Clinical and hemodynamic outcomes for transcatheter aortic valve implantation in internally stented surgical valve -Comparison of Balloon- and Self-Expandable Valves-

**DOI:** 10.1101/2025.06.24.25330238

**Authors:** Daisuke Sato, Noriaki Moriyama, Yoichi Sugiyama, Hirokazu Miyashita, Tomoki Ochiai, Koki Shishido, Futoshi Yamanaka, Tommi Vähäsilta, Teemu Laakso, Sebastian Dahlbacka, Tiina Vainikka, Juho Viikilä, Shigeru Saito, Mika Laine, Mikko Jalanko

**Affiliations:** Department of Cardiology, Heart and Lung Center, Helsinki University Hospital and University of Helsinki, Helsinki, Finland; Department of Cardiology, Shonan Kamakura General Hospital, Kanagawa, Japan; Department of Cardiovascular surgery, Heart and Lung Center, Helsinki University Hospital and University of Helsinki, Helsinki, Finland

**Keywords:** transcatheter aortic valve replacement, internally stented valves, Mitroflow, Trifecta, balloon-expandable valve, self-expandable valve

## Abstract

**Background:** Valve-in-valve (ViV) transcatheter aortic valve implantation (TAVI) is indicated in patients undergoing repeat interventions for degenerative aortic valve bioprostheses. Patients with internally stented surgical valves (IS) (Mitroflow and Trifecta) are at a high risk for coronary artery obstruction during the ViV procedure. This study aimed to evaluate the mid-term clinical and hemodynamic outcomes of balloon-expandable valves (BEV) and supra-annular self-expanding valves (SEV) for TAVI within the IS.

**Methods:** Baseline characteristics, hemodynamic parameters, and clinical outcomes of patients who underwent ViV for IS treated with BEV and SEV were retrospectively collected. Outcomes were compared using propensity score matching (PSM).

**Results:** In total, 113 patients were included this analysis. Sixty-three patients (55.8%) underwent BEV, and fifty patients (44.2%) underwent SEV. Overall, 37 pairs were identified after PSM. At 30-day, the clinical and hemodynamic outcomes were similar between the groups. Patients with SEV had better post-procedural mean gradient at 1-year compared with those with BEV (22.7±8.4 mmHg vs 11.7±4.3 mmHg; p<0.001). There was no significant difference between SEV and BEV in the cumulative 2-year composite endpoint, including all-cause of mortality, hospitalization for heart failure, and coronary obstruction (log-rank p=0.489)

**Conclusions:** In patients who underwent ViV for IS, the early- and mid-term clinical outcomes were comparable between BEV and SEV. Meanwhile, the 1-year hemodynamics seemed to be better in patients with SEV than in those with BEV.

**Clinical Perspective:** What is new?

- Mid-term clinical outcomes, including all-cause mortality, heart failure rehospitalization, and coronary obstruction, were similar between balloon-expandable valve (BEV) and self-expanding valve (SEV) in valve-in-valve transcatheter aortic valve implantation (ViV-TAVI) for internally stented surgical valves (IS).
- SEV group has lower transvalvular gradients at 1-yearfollow-up than BEV group, while the occurrence of coronary obstruction were similar.

What are the Clinical Implications?

- Individualized device selection and improving pre-operative screening for a risk of coronary obstruction, patient-specific anatomical and procedural factors can help the better clinical outcomes.

## Introduction

Valve-in-valve (ViV) transcatheter aortic valve implantation (TAVI) is a well-established alternative treatment option for patients with degenerated surgical bioprostheses who are at high surgical risk for redo surgical aortic valve replacement.^1–2^ Among these surgical aortic valves, internally stented valves (IS) (Mitroflow [Sorin Group] and Trifecta [St.Jude Medical]) have unique features, in which pericardial leaflets are mounted outside the valve stent. The externally mounted pericardial leaflets of these prostheses may be displaced away from the valve stent during deployment of the TAVI prosthesis, potentially leading to coronary obstruction.^3–4^ In addition, as these valves have often been used for small native annuli, future ViV-TAVI in the small-sized bioprostheses has a high risk of significant residual stenosis.^6^ These factors raise concerns about adverse outcomes and long-term durability in patients undergoing ViV-TAVI.

The presence of a small aortic annulus is associated with poor outcomes and suboptimal hemodynamics after surgical aortic valve replacement, which can lead to patient-prosthesis mismatch (PPM) and high-residual gradient.^2^ Previous studies have shown that in patients with failed small surgical aortic bioprostheses undergoing ViV-TAVI, supra-annular self-expanding transcatheter heart valves (SEV) are associated with lower transvalvular gradient and better valve performance, but not with adverse outcomes, compared with balloon-expandable transcatheter heart valves (BEV). ^2,5^ However, no studies have directly compared the clinical and hemodynamic outcomes of ViV-TAVI within the IS between BEV and SEV. Therefore, this study aimed to compare the clinical and hemodynamic outcomes of BEV and SEV in patients undergoing ViV-TAVI for IS surgical bioprostheses using a propensity score-matched cohort.

## Methods

### Study population

The international TAVI registry (Trial Identifier: UMIN000040413) was a retrospectively and prospectively collected data from consecutive patients who underwent TAVI for severe aortic stenosis (AS) or degenerated surgical aortic valves at two high-volume centers: Helsinki University Hospital in Finland and Shonan Kamakura General Hospital in Japan. A total of 163 patients who underwent ViV-TAVI with SEV or BEV between December 2008 and February 2024 were enrolled in this analysis. We excluded TAVI cases for patients who underwent TAV-in-TAV or TAV-in-without IS. A total of 113 patients who underwent ViV-TAVI for IS were included in this study. Thereafter, a 1:1 propensity score matching (PSM) analysis was conducted, resulting in two paired groups: SEV (n = 36) and BEV (n = 36) groups (Figure 1). The SEVs available during the study period were Evolut R, PRO, PRO+, and FX (Medtronic, Minneapolis, MN, USA), and the BEVs were SAPIEN 3, and Ultra (Edwards Lifesciences, Irvine, CA, USA). Indications for ViV were categorized depending on aortic prosthesis dysfunction as follows: 1) prosthesis stenosis: mean prosthesis gradient ≥ 40 mm Hg (severe AS) with aortic insufficiency (AI) less than moderate; 2) AI: moderate or greater transvalvular AI with mean prosthesis gradient < 40 mm Hg; and 3) mixed: mean prosthesis gradient ≥ 40 mm Hg with moderate or greater transvalvular AI. The prospective registry was established in accordance with the principles of the Declaration of Helsinki and was approved by the Institutional Clinical Research and Ethics Committee. Informed patient consent was waived due to the retrospective nature of the study. The data that support the findings this study is available from the corresponding author upon reasonable request.

**Figure 1.**
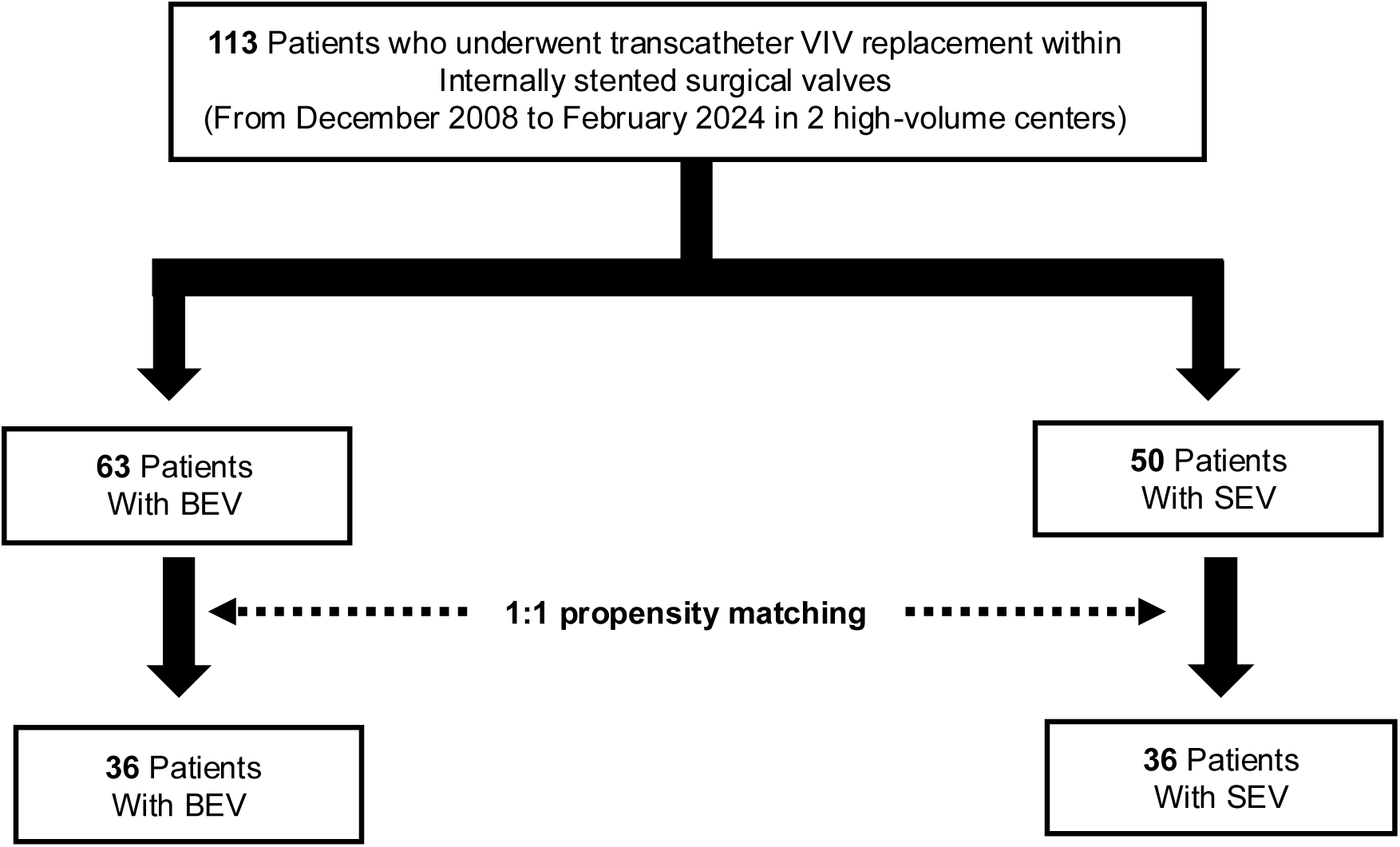
**Study groups flowchart** Study groups flowchart. VIV=valve-in-valve; BEV=balloon expandable valve SEV=self-expanding valve

### Device description and definition

The decision to proceed with ViV-TAVI was made with the consensus of a dedicated heart team, including experienced clinical and interventional cardiologists and cardiovascular surgeons. Transcatheter heart valve (THV) sizing was determined by smartphone application-based sizing data, manufacturer’s tables, 3D multidetector-row CT-based (MDCT) annular measurements, and expert consensus.^6^ Valve sizes of the surgical valves were defined on the basis of true internal diameter (ID).^7^ After the procedure, the patients were evaluated for postprocedural hemodynamics and cardiac function using transthoracic echocardiography (TTE) during hospitalization. Clinical outcomes and procedural complications such as stroke, bleeding, vascular complications, acute kidney injury, and newly required pacemaker implantation were analyzed according to the Valve Academic Research Consortium 3 criteria.^8^

### Coronary protection and risk for occlusion

Coronary protection was performed if there was potential risk for occlusion depending on generally published data^9^ (virtual valve-to-coronary distance ≤4 mm, height of the coronary ostium ≤10 mm, and shallow sinus of Valsalva average diameter ≤30 mm). It was also performed if there was specific patient anatomy (whether the presence of leaflet calcium, which valve is anticipated to be used [BEV versus SEV]). If there are high-risk features for coronary obstruction, coronary protection is performed before deploying the valve by engaging the coronary ostium with an appropriately sized stent.

### Data collection and outcomes

The baseline clinical, procedural, and follow-up data were prospectively recorded in a database maintained by each institute. Adverse events were systematically recorded and adjudicated by an independent clinical event committee. Data were collected from a collaborative registry database. Clinical follow-up data were obtained by reviewing medical records or telephone interviews.

The primary endpoints were 30-day and 2-year clinical outcomes, including all-cause death, hospitalization for heart failure, and coronary obstruction. Secondary outcomes were hemodynamics evaluated using TTE at 30-day and 1-year.

### Statistical analysis

Categorical variables were described as absolute numbers with relative percentages and compared using the X^2^ or Fisher’s exact test, as appropriate. Continuous variables were expressed as mean±standard deviation (SD) or median (interquartile range) and compared using the Student’s t-test or the Mann-Whitney U test depending on the distribution of the variables. Given the differences in baseline characteristics and selection bias between the SEV and BEV groups, PSM analysis was applied to identify a cohort of patients with similar baseline characteristics. The PSM was performed on the basis on a logistic regression model constructed with the following variables: age, sex, body mass index (BMI), Society of Thoracic Surgeons (STS) mortality score, Euro SCORE2, hypertension, diabetes mellitus, atrial fibrillation, chronic kidney disease, left ventricular ejection fraction, chronic pulmonary disease, peripheral artery disease, and true ID. Patients were matched using the PSM logit with 1:1 optimal matching and a caliper width of 0.2. The cumulative incidence of clinical events was estimated using the Kaplan-Meier method, and differences were assessed using the log-rank test. Multivariate COX regression analysis was performed to identify independent predictors of the composite endpoints, including all-cause mortality, hospitalization for heart failure, and coronary obstruction at 2 years. Clinically relevant baseline and procedural variables were used with p values <0.20 in the univariate analysis. A two-sided p value < 0.05 was considered statistically significant for all analyses. All analyses were carried out using SPSS version 29 (IBM, Armonk, NY).

## Results

### Baseline patient characteristics

During the study period, a total of 113 patients were included in this analysis. 63 patients were treated with BEV and 50 patients with SEV. The baseline characteristics of the study cohort are shown in **Table 1**. The SEV group included more women (49.2% vs 72.0%, p=0.014) compared with BEV group. The BEV group had a higher proportion of chronic pulmonary disease, and peripheral artery disease than the SEV group (p=<0.01 and p=0.011, respectively). Prior percutaneous coronary intervention and coronary artery bypass grafting (CABG) were not different between the two groups. The median time between the index surgical aortic valve replacement and the ViV-TAVI procedure was similar in both groups. The SEV group had a higher proportion of aortic stenosis than the BEV group (p=0.035). The PSM resulted in 36 matched pairs. After the matching, the absolute standardized mean difference (d-value) of all covariates was ≤0.2. The patient’s baseline characteristics were well balanced between the BEV and SEV groups, including TTE and MDCT variables.

**Table 1.**
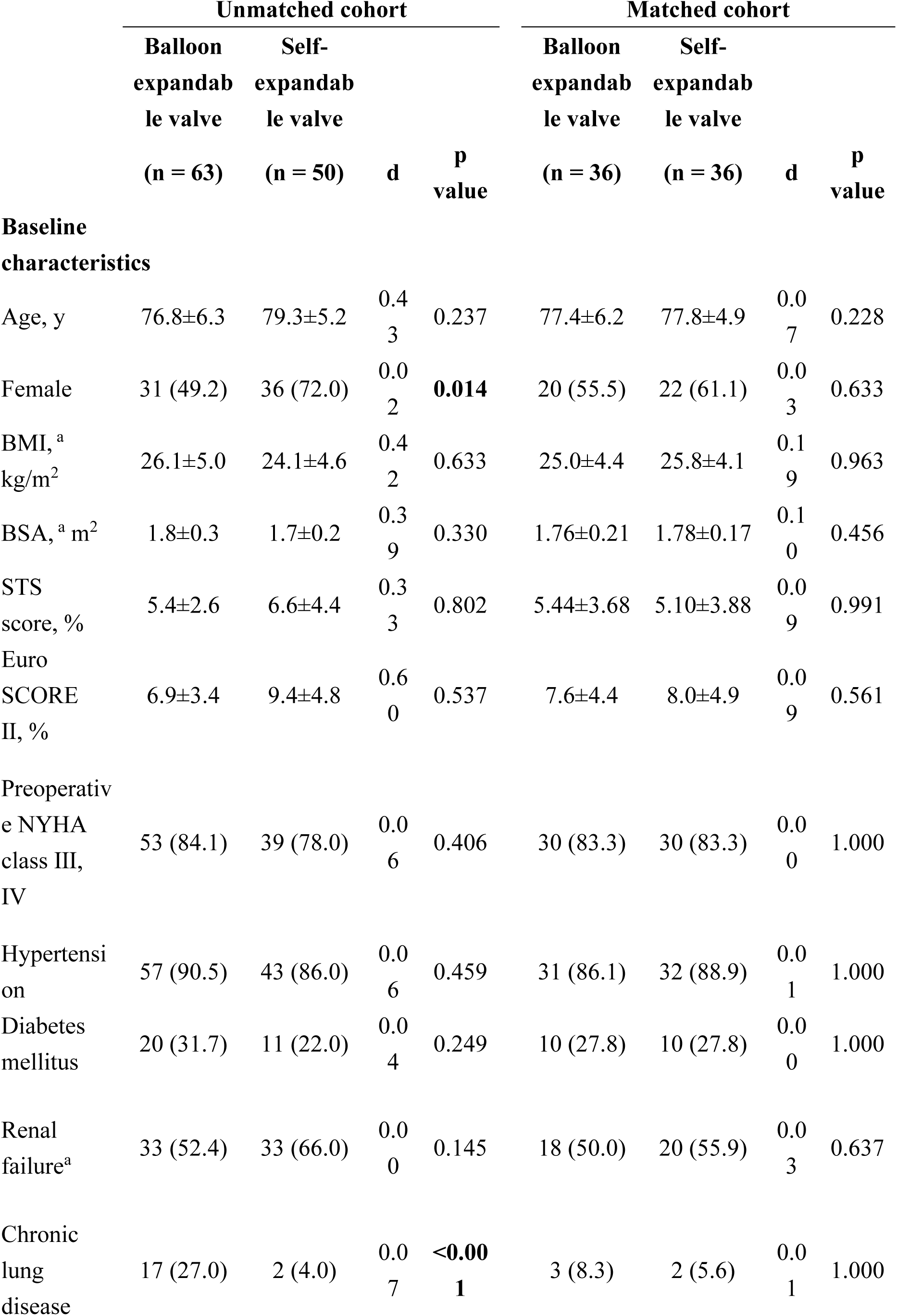
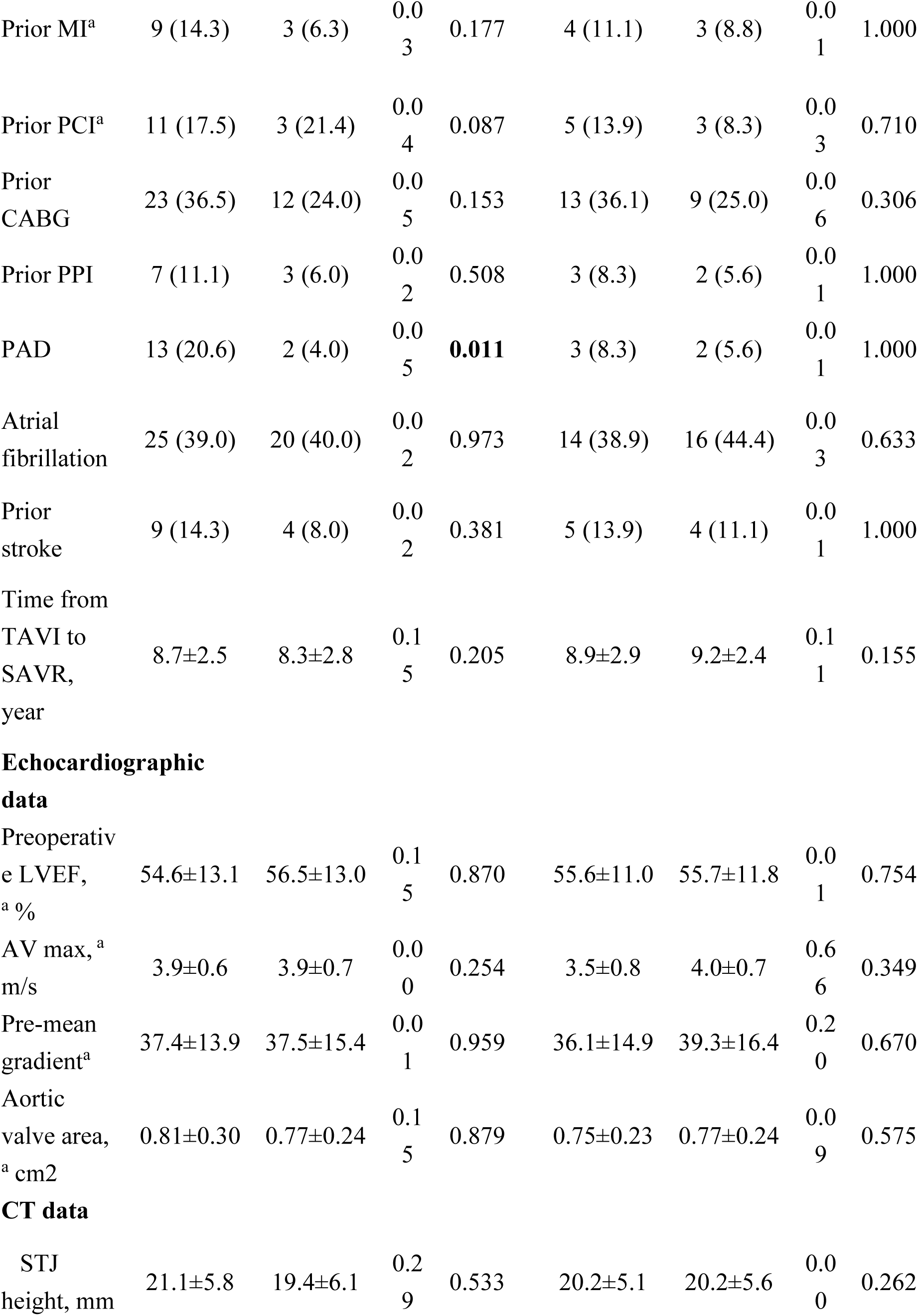
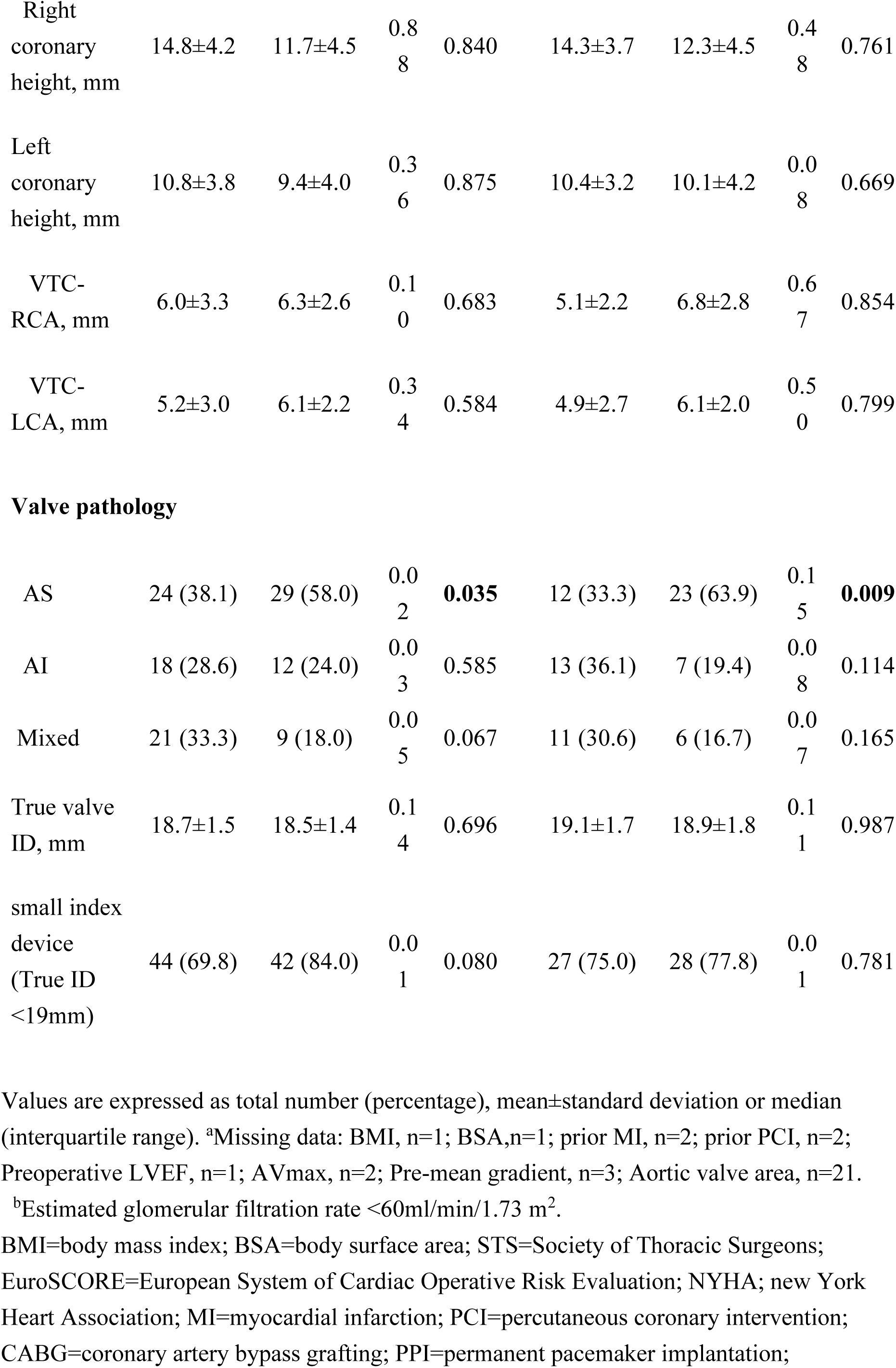
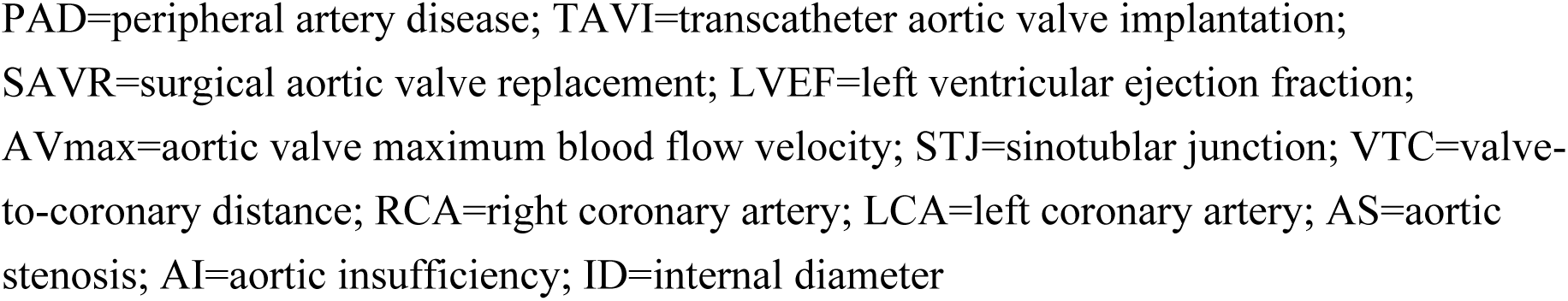
Baseline characteristics.

### Procedural outcomes

The procedural outcomes are summarized in **Table 2**. As the results were consistent between the unmatched and matched cohorts, only the findings for the matched cohorts are presented. Most patients underwent TAVI via a transfemoral approach. Coronary protection against potential coronary obstruction and required chimney stenting deployment were similar between the two groups (13.9% vs 11.1%; p=1.000, 11.1% vs 11.1%; p=1.000, respectively). One patient in the BEV group occurred acute coronary obstruction owing to sinus sequestration (**Supplement Figure 1**). In the SEV group, post-dilatation was performed more frequently than in the BEV group (2.8% vs 25.0%; p=0.014). There were no significant differences in the mean pressure gradient after TAVI. No patients in either group had moderate or severe aortic regurgitation (AR). The incidence of mild AR was higher in the SEV group than in the BEV group (5.6% vs 17.1%).

**Table 2.**
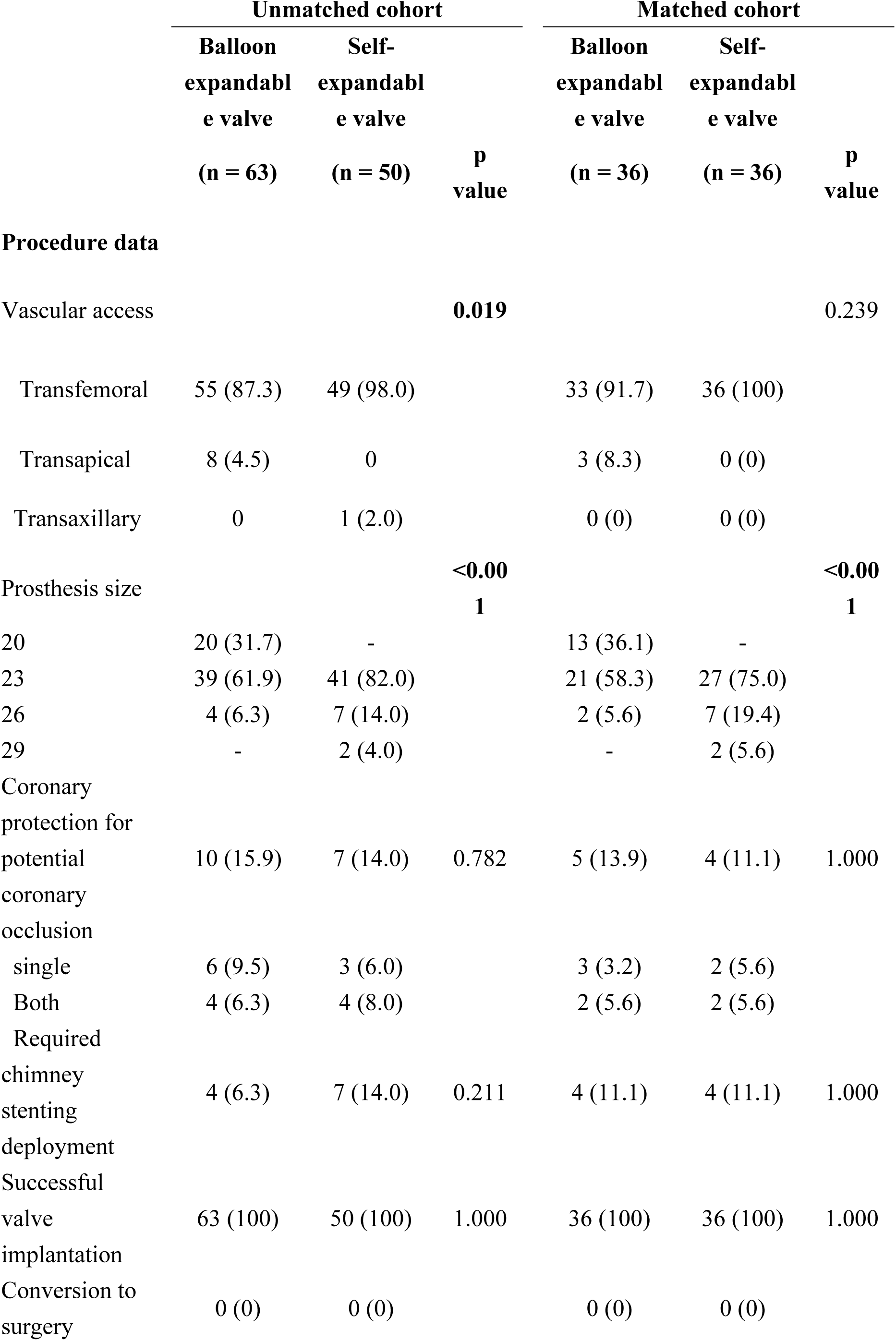
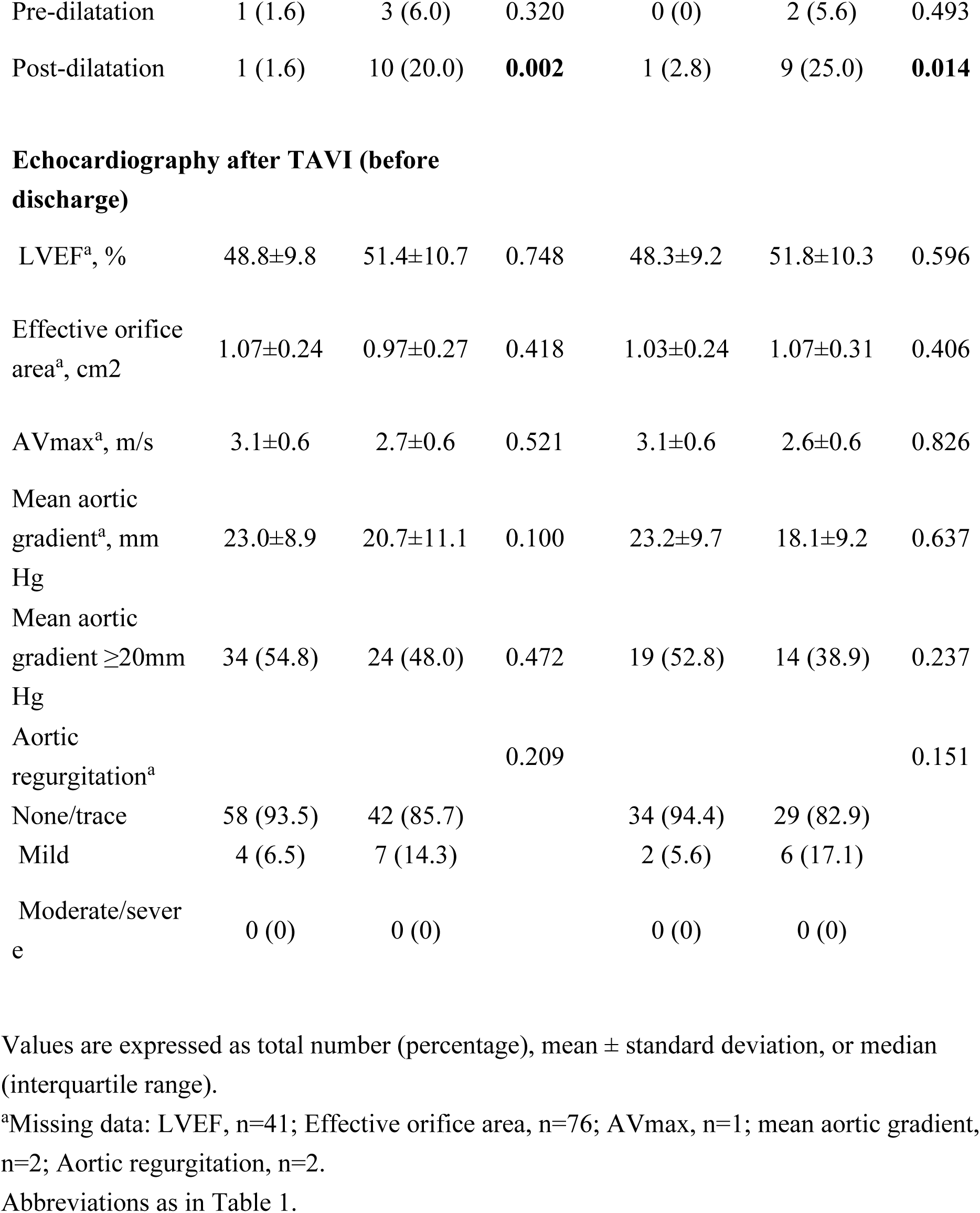
Procedural results.

|  | Unmatched cohort |  |  | Matched cohort |  |  |
| --- | --- | --- | --- | --- | --- | --- |
|  | Balloon expandable valve<br>(n = 63) | Self-expandable valve<br>(n = 50) | P value | Balloon expandable valve<br>(n = 36) | Self-expandable valve<br>(n = 36) | P value |
| **Procedure data** |  |  |  |  |  |  |
| Vascular access |  |  | **0.019** |  |  | 0.239 |
| Transfemoral | 55 (87.3) | 49 (98.0) |  | 33 (91.7) | 36 (100) |  |
| Transapical | 8 (4.5) | 0 |  | 3 (8.3) | 0 (0) |  |
| Transaxillary | 0 | 1 (2.0) |  | 0 (0) | 0 (0) |  |
| Prosthesis size |  |  | **<0.001** |  |  | **<0.001** |
| 20 | 20 (31.7) | - |  | 13 (36.1) | - |  |
| 23 | 39 (61.9) | 41 (82.0) |  | 21 (58.3) | 27 (75.0) |  |
| 26 | 4 (6.3) | 7 (14.0) |  | 2 (5.6) | 7 (19.4) |  |
| 29 | - | 2 (4.0) |  | - | 2 (5.6) |  |
| Coronary protection for potential coronary occlusion | 10 (15.9) | 7 (14.0) | 0.782 | 5 (13.9) | 4 (11.1) | 1.000 |
| single | 6 (9.5) | 3 (6.0) |  | 3 (3.2) | 2 (5.6) |  |
| Both | 4 (6.3) | 4 (8.0) |  | 2 (5.6) | 2 (5.6) |  |
| Required chimney stenting deployment | 4 (6.3) | 7 (14.0) | 0.211 | 4 (11.1) | 4 (11.1) | 1.000 |
| Successful valve implantation | 63 (100) | 50 (100) | 1.000 | 36 (100) | 36 (100) | 1.000 |
| Conversion to surgery | 0 (0) | 0 (0) |  | 0 (0) | 0 (0) |  |
| Pre-dilatation | 1 (1.6) | 3 (6.0) | 0.320 | 0 (0) | 2 (5.6) | 0.493 |
| Post-dilatation | 1 (1.6) | 10 (20.0) | **0.002** | 1 (2.8) | 9 (25.0) | **0.014** |
| **Echocardiography after TAVI (before discharge)** |  |  |  |  |  |  |
| LVEF<sup>a</sup>, % | 48.8±9.8 | 51.4±10.7 | 0.748 | 48.3±9.2 | 51.8±10.3 | 0.596 |
| Effective orifice area<sup>a</sup>, cm<sup>2</sup> | 1.07±0.24 | 0.97±0.27 | 0.418 | 1.03±0.24 | 1.07±0.31 | 0.406 |
| AVmax<sup>a</sup>, m/s | 3.1±0.6 | 2.7±0.6 | 0.521 | 3.1±0.6 | 2.6±0.6 | 0.826 |
| Mean aortic gradient<sup>a</sup>, mm Hg | 23.0±8.9 | 20.7±11.1 | 0.100 | 23.2±9.7 | 18.1±9.2 | 0.637 |
| Mean aortic gradient ≥20mm Hg | 34 (54.8) | 24 (48.0) | 0.472 | 19 (52.8) | 14 (38.9) | 0.237 |
| Aortic regurgitation<sup>a</sup> |  |  | 0.209 |  |  | 0.151 |
| None/trace | 58 (93.5) | 42 (85.7) |  | 34 (94.4) | 29 (82.9) |  |
| Mild | 4 (6.5) | 7 (14.3) |  | 2 (5.6) | 6 (17.1) |  |
| Moderate/severe | 0 (0) | 0 (0) |  | 0 (0) | 0 (0) |  |

Values are expressed as total number (percentage), mean ± standard deviation, or median (interquartile range).
<sup>a</sup>Missing data: LVEF, n=41; Effective orifice area, n=76; AVmax, n=1; mean aortic gradient, n=2; Aortic regurgitation, n=2.
Abbreviations as in Table 1.

### Clinical outcomes and follow-up results

Regarding the 30-day clinical outcomes, there were no significant differences between the two groups in terms of adverse events such as death, stroke, permanent pacemaker implantation, acute kidney injury, major vascular complications, and major/life-threatening bleeding (Table 3).

**Table 3.** Follow-up data.

| Unmatched cohort | Matched cohort |
| --- | --- |

|  | Balloon<br>expandabl<br>e valve<br>(n = 63) | Self-<br>expandabl<br>e valve<br>(n = 50) | P<br>value | Balloon<br>expandabl<br>e valve<br>(n = 36) | Self-<br>expandabl<br>e valve<br>(n = 36) | P<br>value |
| --- | --- | --- | --- | --- | --- | --- |
| **At 30-day follow-up** |  |  |  |  |  |  |
| all-cause of death | 2 (3.2) | 1 (2.0) | 1.000 | 1 (2.8) | 0 (0) | 1.000 |
| Cardiovascular death | 2 (3.2) | 1 (2.0) | 1.000 | 1 (2.8) | 0 (0) | 1.000 |
| Stroke | 0 (0) | 0 (0) |  | 0 (0) | 0 (0) |  |
| Acute Coronary artery occlusion | 1 (1.6) | 0 (0) | 1.000 | 1 (2.8) | 0 (0) | 1.000 |
| Permanent Pacemaker<sup>a</sup> | 1 (1.6) | 2 (3.2) | 0.583 | 0 (0) | 2 (5.6) | 0.493 |
| acute kidney injury | 2 (3.2) | 2 (3.2) | 1.000 | 1 (2.8) | 1 (2.8) | 1.000 |
| VCD success<sup>b</sup> | 54 (98.2) | 47 (95.9) | 0.600 | 32 (97.0) | 35 (97.2) | 1.000 |
| Major/life-threatening bleeding | 9 (14.3) | 6 (12.0) | 0.722 | 4 (11.1) | 4 (11.1) | 1.000 |
| Valve reintervention | 0 (0) | 0 (0) |  | 0 (0) | 0 (0) |  |
| **At 1-year** |  |  |  |  |  |  |
| all cause of death | 6 (9.5) | 4 (8.0) | 1.000 | 3 (8.3) | 3 (8.3) | 1.000 |
| Death or Heart failure hospitalization | 7 (11.1) | 6 (12.0) | 0.883 | 3 (8.3) | 5 (13.9) | 0.710 |
| Late Coronary occlusion | 0 (0) | 0 (0) |  | 0 (0) | 0 (0) |  |
| **Echocardiography at 1-year (n=67)** |  |  |  |  |  |  |
|  | **n=35** | **n=32** |  | **n=18** | **n=22** |  |
| LVEF, <sup>b</sup> % | 58.5±9.5 | 60.9±9.7 | 0.936 | 61.8±7.0 | 59.9±10.0 | 0.165 |
| Mean aortic gradient,<sup>b</sup> mm Hg | 23.4±9.1 | 13.1±6.3 | 0.080 | 22.7±8.4 | 11.7±4.3 | **0.020** |
| Mean aortic gradient ≥20mm Hg<sup>b</sup> | 20 (58.8) | 4 (12.1) | **<0.001** | 11 (61.1) | 1 (4.5) | **<0.001** |
| Aortic regurgitation<sup>b</sup> |  |  | 1.000 |  |  | 1.000 |
| None-trace | 33 (94.3) | 31 (96.9) |  | 17 (94.4) | 21 (95.5) |  |
| Mild | 2 (5.7) | 1 (3.1) |  | 1 (5.6) | 1 (4.5) |  |
| Moderate/severe | 0 (0) | 0 (0) |  | 0 (0) | 0 (0) |  |
| **long-term follow-up (median 1296 days [734-1833])** |  |  |  |  |  |  |
| All cause of death | 21 (33.3) | 10 (20.0) | 0.115 | 8 (22.2) | 8 (22.2) | 1.000 |
| Late coronary occlusion | 0 (0) | 1 (2.0) | 0.442 | 0 (0) | 0 (0) |  |
| Valve reintervention | 1 (1.6) | 0 (0) | 1.000 | 0 (0) | 0 (0) |  |

Values are expressed as total number (percentage), mean ± standard deviation, or median (interquartile range).
<sup>a</sup>Patients with previous PPI were excluded.
<sup>b</sup>Missing data: LVEF, n=42; mean aortic gradient, n=43; Aortic regurgitation, n=42.
VCD=vascular closure device. Other abbreviations as in Table 1.

**Table 4.** Cox Regression Analysis of composite endpoint at 2 years.

|  | **HR**<br>**(95%CI)** | **p value** | **Adjusted HR**<br>**(95%CI)** | **p value** |
| --- | --- | --- | --- | --- |
| Age, pery | 1.02 (0.93-1.12) | 0.648 |  |  |
| Female | 0.52 (0.19-1.40) | 0.195 |  |  |
| BMI | 1.02 (0.92-1.12) | 0.775 |  |  |
| LVEF, % | 0.98 (0.95-1.02) | 0.419 |  |  |
| small valve (True ID<19) | 0.93 (0.30-2.87) | 0.893 |  |  |
| etiology AS | 2.55 (0.89-7.33) | 0.083 | 2.11 (0.71-6.26) | 0.180 |
| etiology AR | 0.64 (0.18-2.25) | 0.489 |  |  |
| etiology mixed | 0.38 (0.09-1.68) | 0.203 |  |  |
| STS score, % | 1.09 (1.05-1.13) | **<0.001** | 1.08 (1.04-1.12) | **<0.001** |
| EURO SCORE II | 1.11 (1.06-1.17) | **<0.001** |  |  |
| Type of THV | 1.00 (0.37-2.68) | 0.998 |  |  |
| Prior PCI | 1.01 (0.23-4.42) | 0.995 |  |  |
| Prior CABG | 0.97 (0.34-2.78) | 0.950 |  |  |
| Prior Permanent<br>Pacemaker | 0.04 (0.00-64.3) | 0.398 |  |  |
| Coronary protection for<br>potential coronary<br>occlusion | 1.88 (0.61-5.84) | 0.273 |  |  |
| Pre-dilatation | 1.94 (0.26-14.7) | 0.522 |  |  |
| Post-dilatation | 1.33 (0.30-5.88) | 0.703 |  |  |
| Pre-mean aortic gradient | 1.01 (0.99-1.04) | 0.330 |  |  |

HR=hazard ratio; THV=transcatheter heart valve. Other abbreviations as in Table 1.

The median long-term follow-up duration for the surviving patients was 1258 days (IQR: 756-1725 days). The incidence of all-cause of mortality at 2 years, and the composite endpoints of all-cause mortality, hospitalization for heart failure, or coronary obstruction at 2 years were similar between the groups (log-rank: p=0.695 and p=0.489, respectively) (**Figure 2**). The mean transvalvular gradient at 1 year was higher in the BEV group than compared with in the SEV group (23 [IQR: 19-33] vs 11 [IQR: 7-16]; p<0.001). The proportion of mean transvalvular gradient ≥ 20mm Hg at 1-year follow-up was significantly higher in the BEV group (61.1% vs 4.5%; p<0001). No differences were observed in the left ventricular ejection fraction and AR between the two groups. In the overall cohort, one case with SEV developed late chimney stent occlusion, and one case with BEV required the ViViV-TAVI procedure due to the valve deterioration during follow-up.

**Figure 2.**
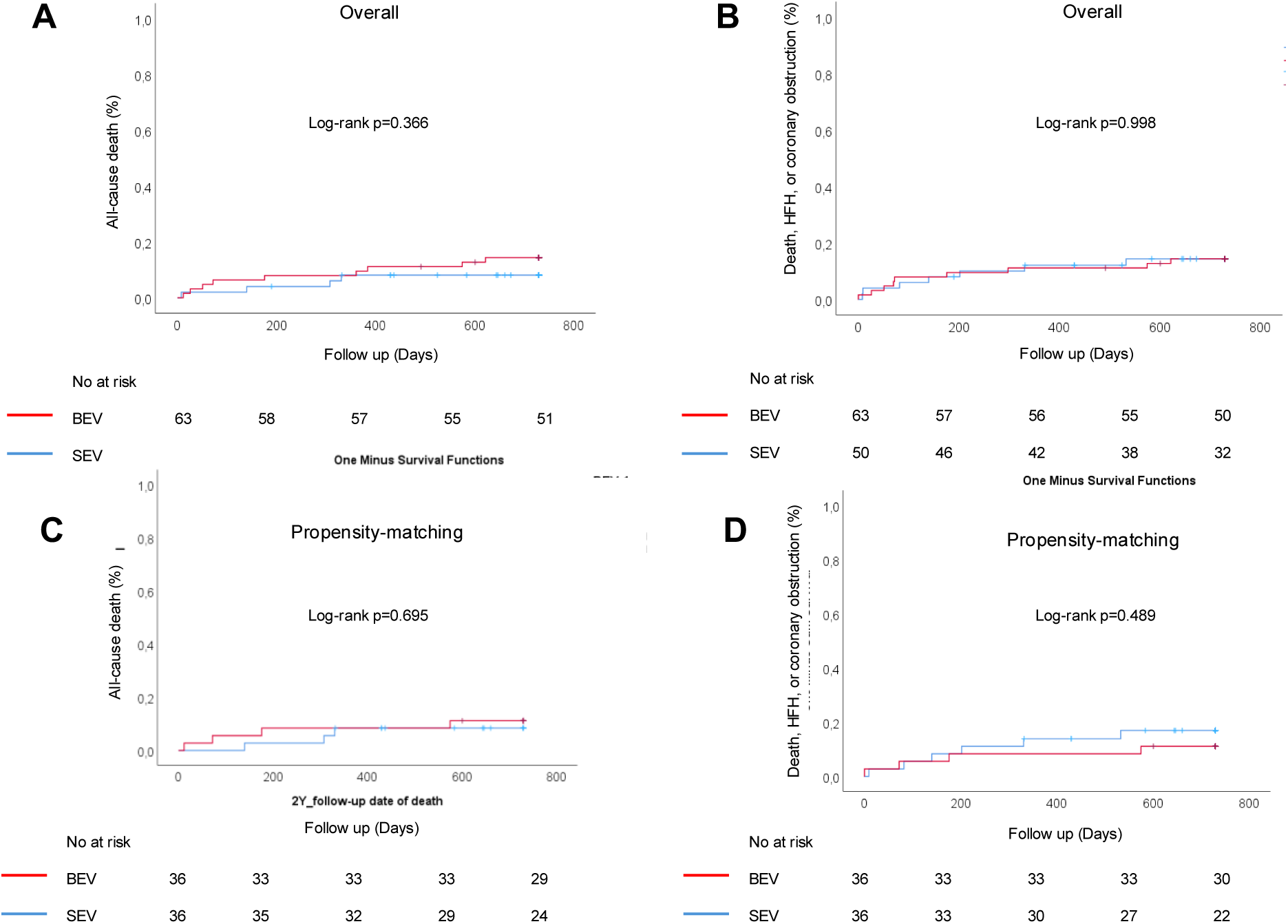
**Cumulative incidence of clinical outcomes** The cumulative incidence of composite outcomes was evaluated using Kaplan-Meier analysis. There was no significant difference between the groups. (A, B) the overall cohort and (C, D) the propensity-matched cohort on the basis of age, sex, BMI, STS mortality score, Euro SCORE2, hypertension, diabetes mellitus, atrial fibrillation, chronic kidney disease, left ventricular ejection fraction, chronic pulmonary disease, peripheral artery disease, and true ID. Abbreviations as in Table 1.

The Cox regression analysis confirmed that STS mortality score (adjusted HR: 1.08; 95% CI: 1.04-1.12; P<0.001) was independently associated with the composite endpoint, including all-cause mortality, hospitalization for heart failure, and coronary obstruction at 2 years.

## Discussion

To the best of our knowledge, this is the first study to compare the clinical and hemodynamic outcomes of BEV and SEV for ViV-TAVI for IS in a PSM population.

The main findings of the current analysis were as follows: in the PSM cohort, 1) early and 2-year clinical outcomes, including all-cause mortality, hospitalization for heart failure, and coronary obstruction, were comparable in patients who underwent VIV-TAVI for IS with BEV and SEV. 2) The SEV group had a lower post-procedural mean transvalvular gradient at the 1-year follow-up than the BEV group.

### Clinical outcomes and hemodynamic status

ViV-TAVI represents an alternative treatment for repeat surgical aortic valve replacement, and the procedure is expected to continue growing in numbers. From the VIVID trial, having a small bioprosthesis represented an important risk factor for mortality after the ViV-TAVI procedure over long-term follow-up.^2^ ViV-TAVI in small surgical bioprostheses has been associated with a higher risk of PPM. Moreover, preexisting PPM has an elevated risk of mortality following ViV TAVI.^2^ In patients who underwent TAVI with native aortic stenosis, especially small annuli, residual transvalvular echocardiographic gradient was lower, and a lower rate of moderate-to-severe PPM was achieved with SEV compared with BEV.^10–12^ This has been partially attributable to the supra-annular design of SEV. Bioprosthetic valve fracture (BVF), which occurs when the surgical bioprosthetic valve ring fractures with a high-pressure balloon inflation, may be an option to improve hemodynamic performance, which result in lower residual transvalvular gradient.^13^ However, BVF has a potential risk for coronary obstruction and THV injury. In our study, about 75% of the patients in the PSM cohort included small index device (True ID ≤ 19mm). The incidence of balloon-dilatation during TAVI procedure was higher in patient with SEV than in those with BEV. Compared with SEV, there was a mean difference of 5 mm Hg higher for the mean transaortic gradient, and a higher proportion of patients having a mean transaortic gradient ≥20 mm Hg before discharge in patients with a BEV. Their differences were maintained at the 1-year follow-up, and the mean transaortic gradient was 11mm Hg higher in patients with a BEV than in those with SEV.

However, the impact of a high residual gradient following TAVI remains inconsistent. High residual transvalvular echocardiographic gradient following ViV-TAVI was not associated with clinical outcomes including major adverse events.^14^ In the randomized LYTEN trial, the supra-annular SEV was associated with lower transvalvular gradient but there were no significant differences in functional status, clinical outcomes, and quality of life at 1-year follow-up, compared with BEV.^5^ In our study, despite a worse hemodynamic profile in patients with BEV, there were no significant differences in composite outcomes of all-cause of mortality, hospitalization for heart failure, or coronary obstruction between the two groups. The Cox regression analysis showed that small valve size (True ID ≤ 19mm), type of THV, and post-dilatation were not the independent predictors of clinical outcomes. This may suggest that the valve type is not associated with mid-term clinical outcomes at 2-years in patients undergoing ViV for IS.

### Coronary protection

Coronary obstruction, including delayed coronary obstruction is a rare but life-threatening complication after ViV-TAVI.^15^ In comparison with other surgical aortic bioprostheses, ViV in IS procedures has a high risk for coronary obstruction due to the stented with externally mounted leaflets bioprostheses.^3^ Compared with the SEV platform, the BEV design presents characteristics of the upper row of open cells and lower stent frame height, which makes it easier to engage coronary cannulation. In our study, one case with BEV had acute coronary obstructions, which contributed to sinus sequestration. Procedural and mid-term outcomes were similar for BEV and SEV, with comparable 2-year composite outcomes. However, one case with SEV had late chimney stent occlusion during follow-up.

Generally, two accepted strategies can be used to prevent coronary obstruction during ViV procedures. First is the chimney/snorkel stenting technique, in which the coronary stent is placed in the coronary artery, and is then implanted in the coronary ostium with protrusion into the aorta after THV implantation if the coronary ostium is occluded. The other is the bioprosthetic aortic scallop intentional laceration to prevent iatrogenic coronary artery obstruction (BASILICA) using transcatheter electrosurgery.^16^ In our study, all patients requiring coronary protection underwent the chimney stenting technique, and most of them required chimney stent implantation. Both chimney stenting and BASILICA effectively prevent TAVI-induced acute coronary obstruction and seem to have acceptable 1-year outcomes.^17^ BASILICA has been introduced to overcome potential limitations of chimney stenting regarding lifetime management, including stent distortion, thrombosis, challenging coronary re-engagement, and stent crushing in case of future valve intervention.^18^

Conversely, BASILICA is a more technically demanding procedure and may not be sufficient to splay the leaflet due to the bulky leaflet and perimeter calcium.^19^ Moreover, long-term follow-up data with these coronary protection techniques were limited. Recently, undermining iatrogenic coronary obstruction with a radiofrequency needle (UNICORN) has been reported as an alternative coronary protection technique for ViV-TAVI procedure.^20^ Further study should be conducted to establish the safety and efficacy of optimal coronary protection strategies.

### Study limitations

First, the study has the inherit limitations of an observational study. Although the current analysis included PSM, the presence of residual confounding factors cannot be ruled out. Second, this study was conducted on a cohort compromising a small number of patients. Therefore, it may have been underpowered to detect differences between the patient groups. Larger cohorts are required to confirm these findings. Third, although the echocardiographic and angiographic parameters were evaluated by experienced cardiologists, our study lacked a core laboratory for imaging analyses. Finally, underreporting or missing echocardiographic and follow-up data need to be acknowledged.

## Conclusions

In patients who underwent ViV-TAVI in the IS group, there were no differences in mid-term clinical outcomes between the SEV and BEV groups. However, those receiving SEV had a lower transvalvular gradient at 1-year follow-up. Further studies with a longer follow-up and larger population will provide information on the impact of SEV and BEV on long-term clinical outcomes, prosthesis durability, and the occurrence of adverse events, including late coronary obstruction and valve reintervention.

## Data Availability

The data that support the findings of this study are available on request from the corresponding author, [NM]

## Acknowledgements

None

## Supplemental Material

Figure S1

## Nonstandard abbreviations and acronyms

ViV: valve-in-valve
TAVI: transcatheter aortic valve implantation
IS: internally stented valves
BEV: balloon-expandable valves
SEV: supra-annular self-expanding valves
AS: aortic stenosis
ID: internal diameter
BMI: body mass index
STS: Society of Thoracic Surgeons
BASILICA: bioprosthetic aortic scallop intentional laceration to prevent iatrogenic coronary artery obstruction

